# High prevalence and risk factors of transmission of hepatitis delta virus in pregnant women in the Center Region of Cameroon

**DOI:** 10.1101/2023.06.08.23291160

**Authors:** Juliette-Laure Ndzie Ondigui, Nadège Mafopa Goumkwa, Cindy Lobe, Brigitte Wandji, Patrick Awoumou, Prisca Voussou Djivida, Puinta Peyonga, Solange Manju Atah, Vivian Verbe, Rachel Kamgaing Simo, Sylvie Agnès Moudourou, Ana Gutierrez, Rosi Garcia, Isabelle Fernandez, Sara Honorine Riwom Essama, Robinson Mbu, Judith Torimiro

**Affiliations:** Department of Microbiology, Faculty of Sciences, University of Yaoundé 1, Yaounde, Cameroon; “Chantal BIYA” International Reference Center for research on HIV/AIDS prevention and management (CIRCB), Yaounde, Cameroon; Department of Food Science and Nutrition, National School of Agro-Industrial Sciences, University of Ngaoundere, Ngaoundere, Cameroon; Yaoundé Gyanecology Obstetrics and Paediatrics Hospital, Yaounde, Cameroon; Department of Biochemistry, Faculty of Medicine and Biomedical Sciences, University of Yaounde 1, Yaounde, Cameroon; Department of Public Health, Faculty of Medicine and Biomedical Sciences, University of Yaounde 1, Yaounde, Cameroon; Cité Verte District Hospital, Yaounde, Cameroon; Biyem-Assi District Hospital, Yaounde, Cameroon; Bikop Catholic Health Center, Bikop, Cameroon; Department of Gynecology/Obstetrics, Faculty of Medicine and Biomedical Sciences, University of Yaounde 1, Yaounde, Cameroon

**Keywords:** risk factors, transmission, hepatitis delta virus, pregnancy

## Abstract

**Background:** Hepatitis B virus (HBV) and hepatitis delta virus (HDV) co-infection has been described as the most severe form of viral hepatitis, and both can be co-transmitted from mother-to-child. A seroprevalence of 4.0% of HDV infection was reported in pregnant women in Yaoundé, and 11.9% in the general population in Cameroon. Our objective was to describe the rate of HDV infection in pregnant women and to determine risk factors associated with mother-to-child transmission of HDV.

**Materials and Methods:** A cross-sectional, descriptive study was conducted from January 2019 to July 2022 among pregnant women attending antenatal contacts in seven health structures in the Centre Region of Cameroon. A consecutive sampling (non-probability sampling) was used to select only women of age over 21 years, who gave a written informed consent. Following an informed consent, an open-ended questionnaire was used for a Knowledge, Attitude and Practice (KAP) survey of these women, and their blood specimens collected and were screened for HBsAg, anti-HIV and anti-HCV antibodies by rapid tests and ELISA. HBsAg-positive samples were further screened for HBeAg, anti-HDV, anti-HBs, and anti HBc antibodies by ELISA, and plasma HDV RNA load measured by RT-qPCR.

**Results:** Of 1992 pregnant women, a rate of 6.7% of HBsAg (134/1992) with highest rate in the rural areas, and 3.9% of hepatitis vaccination rate were recorded. Of 134, 44 (32.3%) were anti-HDV antibody-positive, and 47.6% had detectable RNA viraemia. Two women of 44 anti-HDV-positive cases (4.5%) were co-infected with HBV and HCV, while 5 (11.4%) with HIV and HBV. Multiple deliveries, the presence of tattoos and/or scarifications were significantly statistically associated with the presence of anti-HDV antibodies. Of note, 80% of women with negative HBeAg and positive anti-HBe serological profile, had plasma HDV RNA load of more than log 3.25 (>10.000 copies/ml).

**Conclusion:** These results show an intermediate rate of HDV infection among pregnant women with high level of HDV RNA viremia, which suggest an increased risk of vertical and horizontal co-transmission of HDV.

## Introduction

Hepatitis B virus (HBV) is the aetiologic agent of hepatitis B. The WHO estimates that 296 million people were living with chronic hepatitis B in 2019 with 1.5 million new infections each year[1], whereas Hepatitis delta virus (HDV) affects nearly 5% of people with chronic hepatitis B worldwide[1]. HDV requires the presence of HBV in order to replicate [2-4] in co-infection or super-infection causing the most serious form of chronic viral hepatitis, given its more rapid progression to hepatocellular carcinoma and death [5-8]. It is also worth noting, the low HBV vaccination rate among pregnant women in Cameroon of less than 2.5% [9].

HBV and HDV can be transmitted through skin lesions (injection, scarification, tattooing), contact with infected blood or blood products, but mostly they are co-transmitted through sexual intercourse and from mother-to-child [10]. Therefore, pregnant women are an index population for both vertical and horizontal transmission of HBV and HDV. However, certain practices favour the transmission of HBV in the Centre Region of Cameroon. They include poor accessibility to healthcare, low rate of vaccination, cultural and traditional beliefs, low rate of condom use, high rate of early age of first sexual intercourse, low rate of official marriage, high rate of home delivery, poor link to prevention of mother-to-child transmission (PMTCT) activities, and to other maternal-infant disease prevention efforts in Cameroon.

HBV/HDV co-transmission from mother-to-child has been described [11, 12]. It is an event that seems rare but remains possible and the risk could increase with high level of plasma load of HBV DNA and HDV RNA in the pregnant woman [13-15], in individuals with multiple sexual partners and/or with scarification [16]. Vaccination against hepatitis B is an effective strategy to prevent HDV infection. Therefore, scaling-up the national vaccination programme against hepatitis B in the general population, could lead to a drop in the incidence of hepatitis D worldwide. In 2018, it was reported that 83.3% of pregnant women attending ANC in Yaounde, Cameroon, knew that vaccination against HBV infection is a method of prevention, and 47.1% knew that HBV could be transmitted from mother-to-child. However, only 2.5% had received three doses of the hepatitis B vaccine [9, 17]. In Cameroon the seroprevalence of HBV (HBsAg) in the general population is 11.2%, about two to three times higher than that of the human immunodeficiency virus (HIV) of 3.4% [18, 19]. In 2021, the Cameroon Ministry of Public Health reported a seroprevalence among pregnant women at first trimester of 5.0% in the Centre Region where this study was carried out against 8.63% of national prevalence [20]. In addition, in Central Africa, HDV seroprevalence is estimated at 25.64% in the general population [7], compared to 46.73% in Cameroon in 2018 [21].

Low hepatitis B vaccination coverage in the sexually active population, and lack of recent epidemiological data on HDV in pregnant women, show that the general population in Cameroon is at risk of continuous spread of HBV and HDV from mother-to-child and horizontally. Thus, our specific objectives were to determine the rates of HDV infection and factors associated with risk of HDV infection among pregnant women.

## Materials and methods

### Study design, enrollment procedure and eligibility criteria

A cross-sectional study was conducted from January 2019 to July 2022 in a tertiary hospital, five District hospitals, and one rural Health Center in the Center Region of Cameroon, among pregnant women attending ANC. A total of 1992 pregnant women of age over 21 years were enrolled (Figure 1) from an anticipated 1475 participants deduced from a statistical formula shown below [22] which is commonly used to determine the minimum sample size in medical studies:

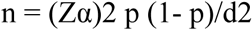

where:

Zα = standard normal variate (1.96 for a 95% confidence interval);

p= prevalence of HDV among pregnant women in Cameroon (4.0) [9];

d = precision of the estimate (0.01).

**Figure 1:**
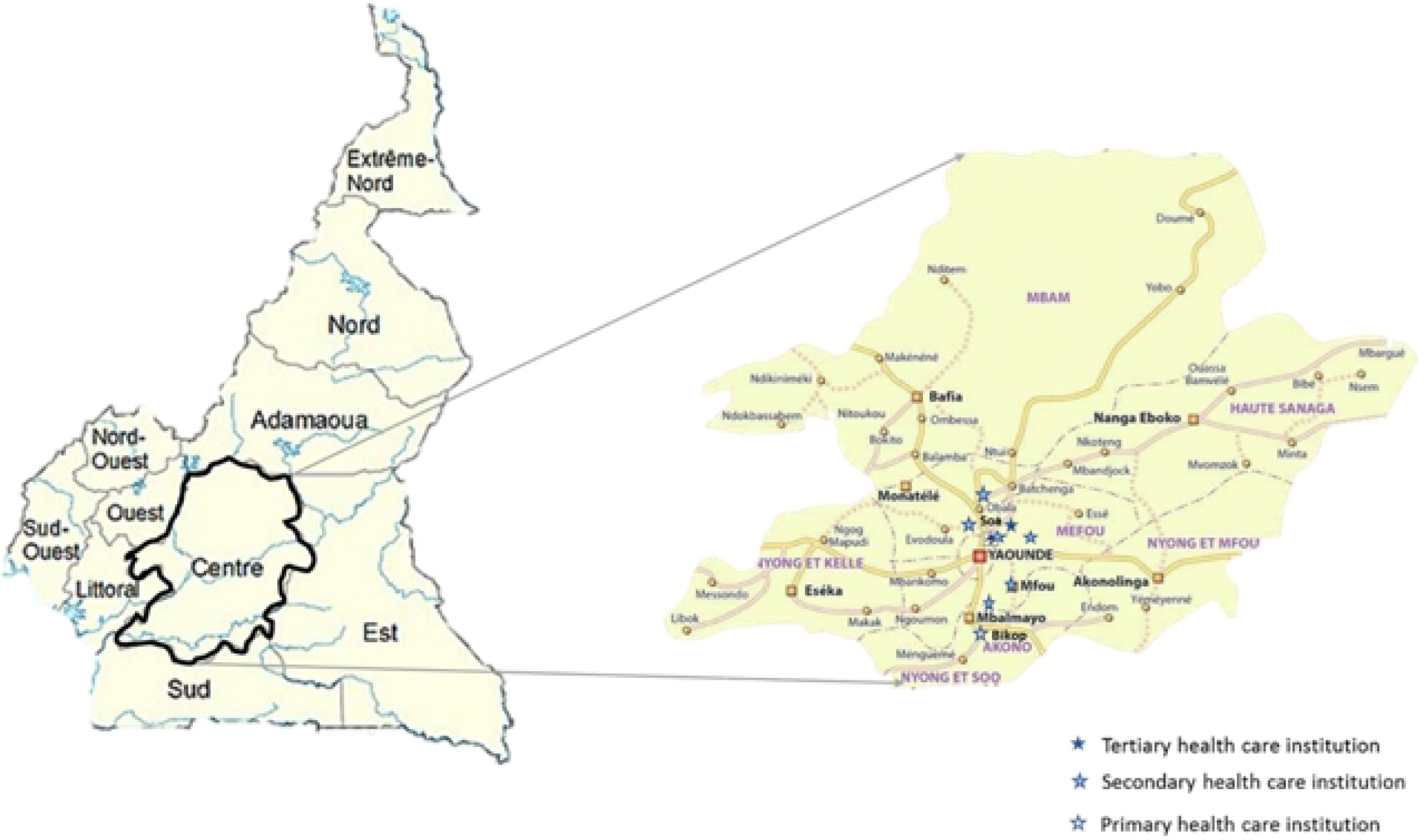
Map of ten regions of Cameroon.

**Figure 2:**
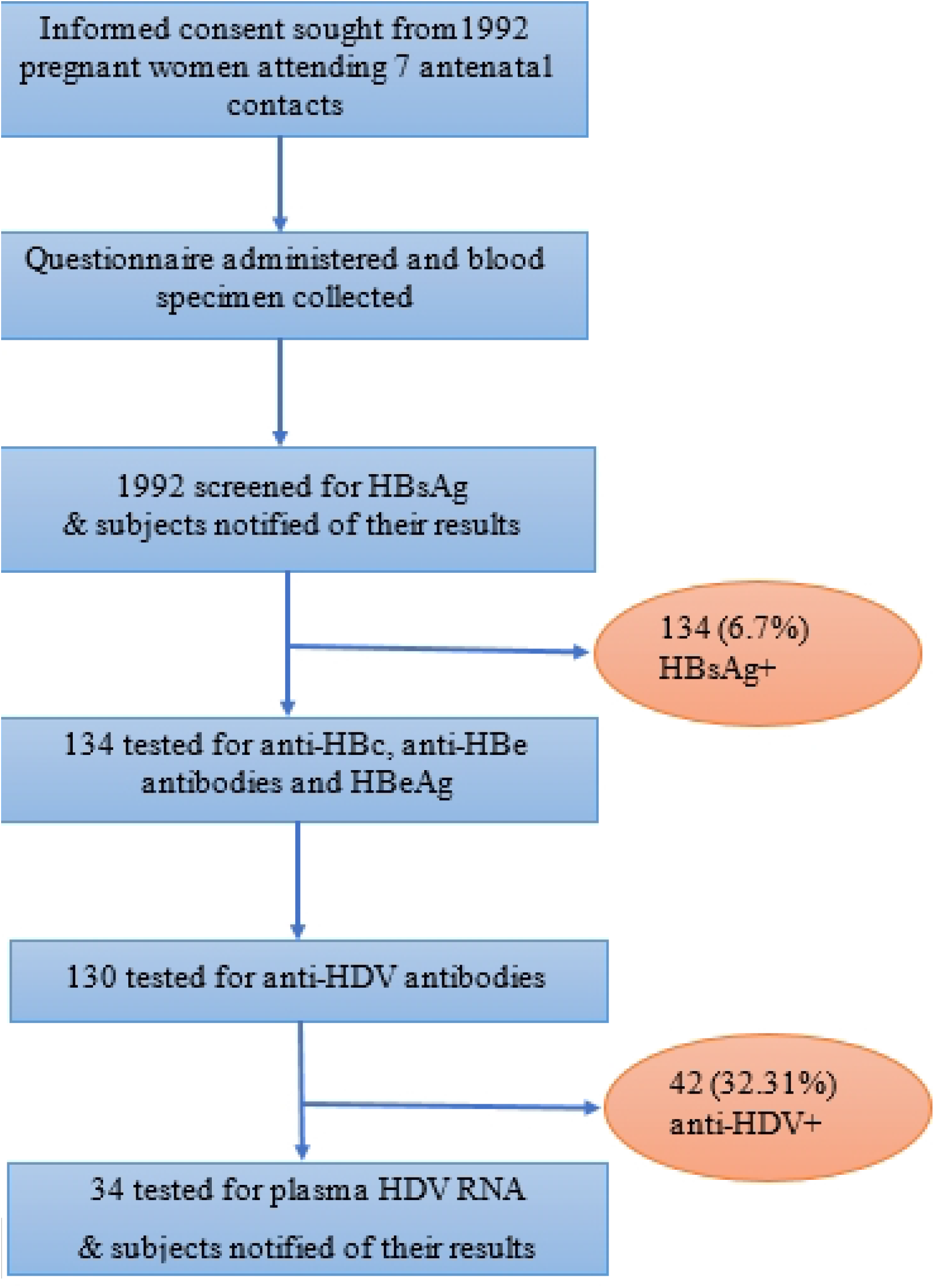
Diagram showing procedure of screening for HBsAg and antibodies to and RNA pf HDV of pregnant women.

Substituting the figures in the above formula yields 1475.2 (nearest whole number). Thus, a minimum of 1475 participants was required.

Data collection took place in 3 stages. Before enrolling the participants, a group sensitization at the hospital or Health Center was conducted to raise awareness on hepatitis B transmission and prevention, hepatitis B and pregnancy, the importance of HBV screening and the purpose of the study. Thereafter, 1992 subjects who were willing to be part of the study, were administered a Consent Form, and upon their consent, a questionnaire with open-ended questions was administered to collect personal and socio-demographic data, their knowledge, attitude and practices with respect to hepatitis B. Some women who took part in the sensitization campaign were not enrolled in the study because they had to have the opinion of their partners, or did not give any reason.

A consecutive sampling (non-probability sampling) was used to select only pregnant women of age over 21 years, who gave a written informed consent. In order to avoid any confusion and to preserve the anonymity of the participants, each participant was assigned a code that allowed us to link each questionnaire to its corresponding blood specimen, while another Register of the ANC used only by hospital staff was established to link the subjects to their results. This same code was used throughout the analysis in the laboratory and the results were linked to the identity of the subject by the hospital staff and not by the research team.

### Laboratory procedures

After completing the questionnaire, 5 ml of blood was drawn from 1992 subjects into a tube containing ethylene-diamine-tetra-acetic acid (EDTA), processed and the plasma tested for HBsAg, anti-HIV and anti-HCV antibodies using rapid diagnostic tests (RDT, ABON, Determine HIV1/2 and KHB Diagnostic kit for HIV antibodies 1 + 2) and ELISA (Murex HBsAg, and Murex HCV Ab), following the manufacturer’s instructions. Furthermore, the HBsAg-positive samples were tested by ELISA for other HBV infection biomarkers: anti-HBs, anti-HBc, HBeAg, and anti-HBe. Positive and negative plasma controls were run alongside each test, following the National Testing Algorithms for these viruses in Cameroon.

ELISA was performed to detect anti-HDV antibodies in 134 plasma samples using the ETI-DELTA-IGMK-2 kit (Dia.Pro Diagnostic Bioprobes srl, Italy). Quantification of HDV RNA (viral RNA load) by real-time qRT-PCR (Eurobioplex HDV Kit, Eurobio Scientific, France) was performed using 34 anti-HDV positive plasma samples. All serologic results were made available to the women within two weeks after specimen collection. The rapid test results were confirmed by two technicians and the samples with borderline results, were further analyzed by ELISA. On the other hand, plasma HDV RNA load was quantified by qRT-PCR commercial kit that detects all 8 main genotypes of HDV, with a lower limit of detection of 100IU/mL. Through these analyses, the subjects that were infected with HBV and/or HDV, were identified. These results were linked to the reported answers given by each subject in relation to their knowledge on modes of transmission and prevention of HBV, their attitude towards prevention of HBV infection such as vaccination and unprotected sex, as well as their practices that predisposes them to acquiring HBV and/or HDV.

All women who tested positive for HBsAg were offered post-test counselling on their status, modes of hepatitis B transmission, need for testing of close contacts for hepatitis B, and vaccination of baby within the first twenty four hours of life against hepatitis B. They were then referred for clinical evaluation and management to a gastroenterologist, and those who were negative for HBsAg were advised to get vaccinated if not yet.

### Statistical analysis

Data collected was regularly checked, corrected, recorded and processed using IBM.SPSS. Statistics.v20 software, while ensuring anonymity with the use of subject registration code. Frequencies of qualitative data were estimated as means. Using internal negative and positive controls in each run, a cycle threshold (Ct) of ≤ 35 was considered positive, and Ct > 35 and ≤ 45 was considered a non-quantifiable positive result. Using a standard curve drawn from standards provided by the manufacturer, the HDV RNA load was extrapolated to copies/mL which was then multiplied by a factor (0.18) to convert into IU/mL. The analysis of the relationship between the prevalence and the different characteristics of the sample was done by logistic regression. Using the Pearson Chi-2 test, the factors associated with HBV and HDV infection, were determined with p values <0.05 considered statistically significant. Ten samples with missing data were not included in the analysis.

### Ethical considerations

The study was conducted in accordance with the fundamental principles of the Declaration of Helsinki, with administrative and ethical approval from health facilities and laboratories, as well as the National Ethics Committee for Human Health Research in Cameroon (N° 2018/06/1150/L/CNERSH/SP), respectively. A written informed consent was obtained from all participants, data was recorded using unique identifiers to ensure confidentiality, and laboratory results were returned to participants for potential benefits in their clinical management. The subjects positive for HBsAg antigen were referred to a gastroenterologist and those HBsAg-negative were advised to be vaccinated if not yet.

## Results

### Socio-demographic parameters and Knowledge, Attitude and Practices of pregnant women

Overall, 1992 pregnant women were tested for HBsAg, with a mean age of 27.53 ± 5.68 years. The average gestational age was 17 ± 6 weeks, with a majority of the women at the second trimester pregnancy. One thousand nine hundred and thirteen (1913) women have never received the hepatitis B vaccine, but 56 had received three doses of the vaccine (2.8%) (Table 1). One thousand six hundred and twenty seven (1627) women were screened for the first time for HBsAg during this study (81.7%). In the group of HBsAg-positive women, 53(39.9%) women were in their first pregnancy, 75 (56.4%) lived with a partner, 37 (27.8%) knew their partner’s hepatitis B status and 5.4% (2/37) of their partners were positive for HBsAg, 15 (11.3%) had already been tattooed or scarified and 21 (15.7%) had a history of surgery.

**Table 1:** Characteristics of the study population.

| Variables |  | HBsAg |  | p-value |
| --- | --- | --- | --- | --- |
|  |  | Negative (%) | Positive (%) |  |
| **Age (years)**<br>**Mean = 27 years** | 15-20 | 209 (91.2) | 19 (8.8) | 0,21 |
|  | 21-25 | 580 (92.2) | 49 (7.8) |  |
|  | 26-30 | 507 (94.9) | 27 (5.1) |  |
|  | 31-35 | 360 (93.0) | 27 (7.0) |  |
|  | 36-40 | 172 (94.0) | 11 (6.0) |  |
|  | 41-45 | 31 (100.0) | 0 (0) |  |
| **Gestational age (weeks)**<br>**Mean = 18 weeks** | 1-14 | 527(92.1) | 45(7.9) | **0 .017\*** |
|  | 15-28 | 940(94.4) | 65(5.6) |  |
|  | 29-42 | 374(90.1) | 41(9.9) |  |
| **Hepatitis B vaccination** | Not vaccinated | 1779(93.04%) | 133(7.0) | 0.637 |
|  | 1 Dose | 10(100.0%) | 0(0) |  |
|  | 2 Doses | 14(100.0%) | 0(0) |  |
|  | 3 Doses | 56(0.58%) | 0(0) |  |
| **Location of health institution/study site** | urban | 1169(94.88%) | 63(5.1) | **0 .016\*** |
|  | semi-urban | 263(91.96%) | **23(8.0)** |  |
|  | rural | 427(90.08%) | **47(9.9)** |  |

\*.The Chi-square statistic is significant at the .05 level.

### Prevalence of HDV and HBV Infection markers

Of 1992 pregnant women tested for HBsAg, 133 were positive (6.7%) while 9.8% of women in the third trimester were HBsAg-positive, and 9.9% of them live in the rural area. Of these, 42 samples were positive for anti-HDV antibodies by serology, with 62.5% with detectable plasma HDV RNA. Meanwhile, rates of 9.0%, 79.7% and 94.0% for HBeAg, anti-HBe and anti-HBc, respectively, was recorded for the HBsAg-positive specimens. Table 2 shows the distribution of serological markers of hepatitis B infection (HBeAg, HBeAb and HBcAb) as a function of the presence of anti-HDV antibodies, indicating that HBeAg was statistically significantly associated with the presence of anti-HDV antibodies (p = 0.008). Two (4.8%) women were co-infected with HCV, HBV and HDV, and 5 (11.9%) with HIV, HBV and HDV. However, HBV/HIV and HBV/HCV co-infections were not associated with anti-HDV positivity in this study. In univariate analysis, neither of HIV nor HCV serologic positivity was associated statistically significantly with the presence of anti-HDV antibodies (Table 2).

**Table 2:** Distribution of viral infections (HCV, HDV, and HIV) and serological markers of hepatitis B infection among HBsAg-positive pregnant women.

| Serological markers |  | Presence of anti-HDV antibodies |  |  |
| --- | --- | --- | --- | --- |
|  |  | Negative (%) | Positive (%) | P-value |
| **Anti-HIV** | Negative | 80(68.4) | 37(31.6) | 0.612 |
|  | Positive | 8(61.5) | 5(38.46) |  |
| **Anti-HCV** | Negative | 88(68.8) | 40(31.2) | 0.119 |
|  | Positive | 0(0) | 2(100.0) |  |
| **HBeAg** | Negative | 82(69.5) | 36(30.5) | 0.169 |
|  | Positive | 6(50.0) | 6(50.0) |  |
| **HBc antibodies** | Negative | 2(25.0) | 6(75.0) | **0.008\*** |
|  | Positive | 86(70.5) | 36(29.5) |  |
| **HBe antibodies** | Negative | 9(36.0) | 16(64.0) | **0.001\*** |
|  | Positive | 79(75.2) | 26(24.8) |  |
| **HBeAg-/HBeAb -** |  | 8(44.4) | 10(55.6) | **0.005\*** |
| **HBeAg+/HBeAb -** |  | 0(0) | 6(100.0) |  |
| **HBeAg-/HBeAb +** |  | 75(75.3) | 26(25.7) |  |
| **HBeAg+/HBeAb +** |  | 5(100.0) | 0(0) |  |

*Note*
✚: positive;
✚: negative;
\*.The Chi-square statistic is significant at the .05 level.

### Factors associated with hepatitis delta virus infection

Our study shows that 4 women with more than 5 children were positive for anti-HDV antibodies. In addition, women living with a partner (44.0%) were more exposed than women living alone (16.4%) to be infected with HDV (p=0.001), while 60.0% of women who had been tattooed or scarified had anti-HDV antibodies (p=0.015) (Table 3). Comparing the seroprevalence of HDV antibodies against biomarkers of HBV infection, there was a statistically significant difference with the presence of anti-HBc (p=0.008), anti-HBe (p=0.001) and HBeAg-negative/Anti-HBe+ (p=**0.005)**. Of note, the HBeAg-negative/anti-HBe-positive profile was the most frequently detected amongst those with plasma HDV RNA viral load of more than log 3.25 (10.000 copies/mL) (Table 4).

**Table 3:** Variation in the prevalence of hepatitis delta according to the clinical characteristics of pregnant women.

| Variable |  | Presence of anti-HDV antibodies |  |  |
| --- | --- | --- | --- | --- |
|  |  | Negative (%) | Positive (%) | p-value |
| **Age Group (years)**<br>**Mean = 26 years** | 15-20 | 16(84.2) | 3 (15.8) | 0.50 |
|  | 21-25 | 31 (66.0) | 16 (34.0) |  |
|  | 26-30 | 17 (60.7) | 11 (39.3) |  |
|  | 31-35 | 18 (69.2) | 8 (30.8) |  |
|  | 36-40 | 6 (60.0) | 4 (40.0) |  |
| **Gestational age (weeks)**<br>**Mean : 22 (weeks)** | 1-14 | 18 (69.2) | 8 (30.2) | 0.92 |
|  | 15-28 | 34 (68.0) | 16 (32.0) |  |
|  | 29-42 | 36 (66.7) | 18 (33.3) |  |
| **Number of life births** | 0 | 40 (75.5) | 13 (24.5) | **0.030** |
|  | 1 | 26 (72.2) | 10 (27.8) |  |
|  | 2 | 10 (62.5) | 6 (37.5) |  |
|  | 3 | 11 (55.0) | 9 (45.0) |  |
|  | 4 | 1 (100.0) | 0 (0.0) |  |
|  | 5+ | 0 (0.0) | 4 (100.0) |  |
| **Marital status** | life alone | 46 (83.6) | 9 (16.4) | **0.001\*** |
|  | married life | 42 (56.0) | 33 (44.0) |  |
| **Knowledge of tattoo** | yes | 6 (40.0) | 9 (60.0) | **0.015\*** |
|  | no | 82 (71.3) | 33 (28.7) |  |
| **History of surgical operations** | yes | 19 (90.5) | 2 (9.5) | **0.015\*** |
|  | no | 69 (63.3) | 40 (36.7) |  |
| **Knowledge of female circumcision** | yes | 5 (100.0) | 0 (00.0) | 0.115 |
|  | no | 83 (66.4) | 42 (33.6) |  |
| **Use of alcohol** | yes | 39 (68.4) | 18 (31.6) | 0.87 |
|  | no | 49 (67.1) | 24 (32.9) |  |
| **Aware of hepatitis B status of close people** | no one knows | 2 (100.0) | 0 (00.0) | 0.189 |
|  | husband | 15 (60.0) | 10 (40.0) |  |
|  | family member | 0 (00.0) | 2 (100.0) |  |
|  | friend | 46 (64.8) | 25 (35.2) |  |

*Note*
*\*.The Chi-square statistic is significant at the .05 level.*

**Table 4:**
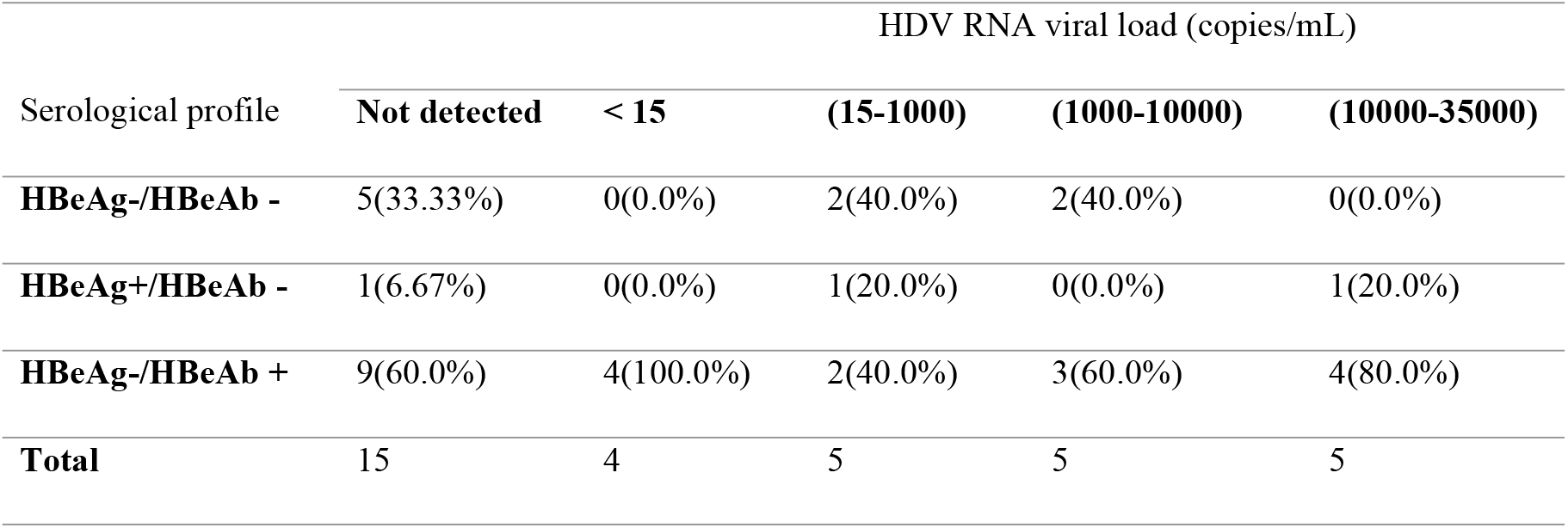
Plasma HDV RNA viral load as a function of the serological profile of HBeAb and HBeAg markers.

## Discussion

We studied the seroprevalence of anti-HDV antibodies and the presence of plasma HDV RNA in 130 HBsAg-positive samples, detected in a sample of 1992 pregnant women collected between January 2019 and July 2022 in seven health structures in the Centre Region of Cameroon. Overall, we observed a seroprevalence of HBsAg of 6.7%. These results are close to those obtained by Torimiro et al in 2018 (6.4%) among pregnant women in Yaoundé, Cameroon [9]. In comparison to other African countries with similar socio-economic and public health contexts, such as Nigeria, Gabon and Burkina Faso, similar prevalence has been reported [23, 24]. We also noted that in our study population the prevalence of HBsAg is higher among pregnant women in rural areas compared to those in urban areas. Other studies conducted in rural areas in Cameroon, have recorded higher prevalence ranging from 7.5% to 13.7% [25-27].

This moderate to high prevalence of hepatitis B in rural areas in Cameroon could be related to the low level of knowledge about hepatitis B modes of transmission and prevention, and risk factors of attitude and practices including idleness and promiscuity. In other words, these are factors that would favour transmission of HDV. Although the efficacy of the hepatitis B vaccine has been proven and has been available since 2005 in the Cameroon EPI [28], this study shows that vaccination coverage remains very low among women of child-bearing age, which contributes in keeping the pool of index infected sub population on the increase. It is therefore not unusual to detect the high prevalence of HDV infection in pregnant women in the Centre Region of Cameroon. Since 1991 when the first study on the prevalence of HDV was conducted in Cameroon[29], several studies have been carried out in the general population or among people with particular socio-demographic characteristics such as pygmies. The variations in the prevalence of anti-HDV antibody ranged from 10.5% to 69% [16, 21, 30-32]. On the other hand, very few of these studies have focused on pregnant women who are an index for both horizontal and vertical transmission of HBV and HDV. These studies reported low levels of anti-HDV antibodies in pregnant women of 2.3-4.0% [9, 29]. These values are low compared to the rate of 32.3% anti-HDV antibody prevalence which we obtained in our study. This difference could be due to the fact that the previous studies were conducted in the Yaoundé an urban area, where there are better living conditions, higher level of education, awareness and knowledge of hepatitis B compared to the semi-urban and rural areas that were included in our study. In comparison of anti-HDV antibody rates among pregnant women in other African countries such as Benin (11.4%), Mauritania (14.7%), Gabon (15.6%), the Central African Republic (21. 4%)[33-36], our result is higher at 32.3%.

Our findings show that the number of children, marital status, presence of tattoos or scarifications and history of surgery are factors associated with the positivity of anti-HDV antibodies. Besombes and colleagues reported in 2020 that individuals with more than 10 sexual casual partners and those who received injections were most at risk of getting HDV [16].

The rates of HBV serological markers in anti-HDV-positive individuals showed that anti-HBc antibodies and anti-HBe antibodies were statistically associated with anti-HDV positivity in this study. Similarly, a majority of women with the HBeAg-negative/HBeAb-positive serological profile, had plasma HDV RNA viral load ranging from 10000 to 35000 copies/mL.

Limitations of this study include the performance of the test HDV antibody test kit and the lack of a national testing algorithm for diagnosis of HDV infection in Cameroon.

## Conclusion

In conclusion, the results of this study show that in spite of the availability of the hepatitis B vaccine in the EPI in Cameroon, HBV infection is moderately endemic (6.7%) in the Centre Region of Cameroon, while HDV rate is 32.3% among pregnant women. Increasing number of casual sexual partners and high level of HDV viremia, may favour horizontal and vertical co-transmission of HDV in this population. Hence, the need to intensify awareness and control strategies for hepatitis B and D among pregnant women and contribute to the global elimination of hepatitis B.

## Data Availability

All relevant data are within the manuscript and its Supporting Information files.

## Acknowledgement

We acknowledge the financial and technical contributions from the Chantal Biya International Reference Centre for Research on Prevention and Management of HIV/AIDS to carry out this study.

## References

1. WHO, Key facts on hepatitis B. 2020.

2. Sawadogo, A., et al., Seroprevalence of hepatitis D virus infection in a population of blood donors carrying HBs Antigen at the Bobo-Dioulasso Regional Blood Transfusion Center. Journal Africain d’Hépato-Gastroentérologie, 2016. 10(1): p. 31–33.

3. Sahin, A., et al., Anti-HDV seroprevalance among patients with previous HBV infection. North Clin Istanb, 2018. 5(2): p. 132–138.

4. Medhat, A., et al., Acute viral hepatitis in pregnancy. Int J Gynaecol Obstet, 1993. 40(1): p. 25–31.

5. Dulger, A.C., et al., High prevalence of chronic hepatitis D virus infection in Eastern Turkey: urbanization of the disease. Arch Med Sci, 2016. 12(2): p. 415–20.

6. A.T. Kabamba, C.M.M., C.M. Nyembo, B.M. Kabamba Et and A.O. Longanga, Hepatitis D seroprevalence among people with chronic hepatitis B in Lubumbashi (DRC). 2020.

7. Stockdale, A.J., et al., Prevalence of hepatitis D virus infection in sub-Saharan Africa: a systematic review and meta-analysis. Lancet Glob Health, 2017. 5(10): p. e992–e1003.

8. Daw, M.A., et al., The Epidemiology of Hepatitis D Virus in North Africa: A Systematic Review and Meta-Analysis. ScientificWorldJournal, 2018. 2018: p. 9312650.

9. Torimiro, J.N., et al., Rates of HBV, HCV, HDV and HIV type 1 among pregnant women and HIV type 1 drug resistance-associated mutations in breastfeeding women on antiretroviral therapy. BMC Pregnancy Childbirth, 2018. 18(1): p. 504.

10. WHO, Key facts on hepatitis D. 2022.

11. Zanetti, A.R., et al., Perinatal transmission of the hepatitis B virus and of the HBV-associated delta agent from mothers to offspring in northern Italy. J Med Virol, 1982. 9(2): p. 139–48.

12. Huang, Y.H., et al., Phylogenetic analysis to document a common source of hepatitis D virus infection in a mother and her child. Zhonghua Yi Xue Za Zhi (Taipei), 1999. 62(1): p. 28–32.

13. Sellier, P., et al., Untreated highly viraemic pregnant women from Asia or sub-Saharan Africa often transmit hepatitis B virus despite serovaccination to newborns. Liver Int, 2015. 35(2): p. 409–16.

14. Sellier, P.O., et al., Hepatitis B Virus-Hepatitis D Virus mother-to-child co-transmission: A retrospective study in a developed country. Liver Int, 2018. 38(4): p. 611–618.

15. Yuan, J., et al., Antepartum immunoprophylaxis of three doses of hepatitis B immunoglobulin is not effective: a single-centre randomized study. J Viral Hepat, 2006. 13(9): p. 597–604.

16. Besombes, C., et al., The epidemiology of hepatitis delta virus infection in Cameroon. Gut, 2020. 69(7): p. 1294–1300.

17. Stockdale, A.J., et al., The global prevalence of hepatitis D virus infection: Systematic review and meta-analysis. J Hepatol, 2020. 73(3): p. 523–532.

18. Bigna, J.J., et al., Seroprevalence of hepatitis B virus infection in Cameroon: a systematic review and meta-analysis. BMJ Open, 2017. 7(6): p. e015298.

19. CAMPHIA 2017 population impact assessment of HIV in Cameroon,. 2020.

20. Directorate for Disease Control, the E.e.l.P.-D., Monitoring Report on Key Health Indicators in Cameroon in 2019. 2022.

21. Butler, E.K., et al., High prevalence of hepatitis delta virus in Cameroon. Sci Rep, 2018. 8(1): p. 11617.

22. Pourhoseingholi, M.A., M. Vahedi, and M. Rahimzadeh, Sample size calculation in medical studies. Gastroenterol Hepatol Bed Bench, 2013. 6(1): p. 14–7.

23. Groc, S., et al., High prevalence and diversity of hepatitis B and hepatitis delta virus in Gabon. J Viral Hepat, 2019. 26(1): p. 170–182.

24. Lingani, M., et al., The changing epidemiology of hepatitis B and C infections in Nanoro, rural Burkina Faso: a random sampling survey. BMC Infect Dis, 2020. 20(1): p. 46.

25. Sone, L.H.E., et al., Prevalence and Identification of Serum Markers Associated with Vertical Transmission of Hepatitis B in Pregnant Women in Yaounde, Cameroon. Int J MCH AIDS, 2017. 6(1): p. 69–74.

26. Eyong, E.M., et al., The prevalence of HBsAg, knowledge and practice of hepatitis B prevention among pregnant women in the Limbe and Muyuka Health Districts of the South West region of Cameroon: a three-year retrospective study. Pan Afr Med J, 2019. 32: p. 122.

27. Mawouma, A.R.N., et al., [Factors associated with hepatitis B infection in pregnant women at health facilities in the health district of Mokolo/Far North of Cameroon]. Pan Afr Med J, 2022. 41: p. 61.

28. Gatchalian, S., et al., A new DTPw-HBV/Hib vaccine is immunogenic and safe when administered according to the EPI (Expanded Programme for Immunization) schedule and following hepatitis B vaccination at birth. Hum Vaccin, 2005. 1(5): p. 198–203.

29. Ndumbe, P.M., Hepatitis D in Yaounde, Cameroon. Apmis, 1991. 99(2): p. 196–8.

30. Noubissi-Jouegouo, L., et al., Evolutionary trends in the prevalence of anti-HDV antibodies among patients positive for HBsAg referred to a national laboratory in Cameroon from 2012 to 2017. BMC Res Notes, 2019. 12(1): p. 417.

31. Luma, H.N., et al., Prevalence and Characteristics of Hepatitis Delta Virus Infection in a Tertiary Hospital Setting in Cameroon. J Clin Exp Hepatol, 2017. 7(4): p. 334–339.

32. Foupouapouognigni, Y., et al., Endemicity and genetic diversity of Hepatitis delta virus among Pygmies in Cameroon, Central Africa. BMC Res Notes, 2022. 15(1): p. 87.

33. Mansour, W., et al., Prevalence, risk factors, and molecular epidemiology of hepatitis B and hepatitis delta virus in pregnant women and in patients in Mauritania. J Med Virol, 2012. 84(8): p. 1186–98.

34. Makuwa, M., et al., Prevalence and genetic diversity of hepatitis B and delta viruses in pregnant women in Gabon: molecular evidence that hepatitis delta virus clade 8 originates from and is endemic in central Africa. J Clin Microbiol, 2008. 46(2): p. 754–6.

35. Komas, N.P., et al., Hepatitis B and hepatitis D virus infections in the Central African Republic, twenty-five years after a fulminant hepatitis outbreak, indicate continuing spread in asymptomatic young adults. PLoS Negl Trop Dis, 2018. 12(4): p. e0006377.

36. De Paschale, M., et al., Prevalence of HBV, HDV, HCV, and HIV infection during pregnancy in northern Benin. J Med Virol, 2014. 86(8): p. 1281–7.

